# Sources of nutrition information used and preferred by older people: A scoping review

**DOI:** 10.1101/2025.02.02.25321554

**Authors:** Jane McClinchy, Angela Dickinson, Emily Barnes, Tai Ibitoye, John Jackson, Amander Wellings

## Abstract

A nutritionally adequate diet is needed for older people to optimise their nutritional status and promote healthy ageing. Making use of nutrition information can inform in people’s food choices to help them improve their health and food experience.

Over a million older people in the UK are at risk of or experience malnutrition. This number is likely to increase as the population ages. Understanding where older people source their nutrition information could help inform public health interventions to reduce the prevalence of malnutrition.

This scoping review aims to summarise studies that examined sources of nutrition information used by older people. A scoping review in order to map and explore studies that examine the current sources of nutritional knowledge used by older people was undertaken using PUBMED, Scopus and CINAHL databases during March 2023. Of the 8819 studies identified, 13 studies reporting on 12 research projects met the inclusion criteria of exposure to, impact by, or preference for at least one source of nutrition information.

Eight studies showed that older people used magazines; 6 studies showed that TV, dietitians, embodied knowledge, family and friends and GPs were the main sources of nutrition information. Educational level, gender and trust in the messages influenced their ability to use these sources to inform their food choices.

Research is needed to further explore the impact of nutritional information in order to identify ways in which to support older adults to make use of nutrition information to help make food choices that would facilitate healthy ageing.

## Introduction

### Malnutrition in older people in the UK

Health eating guidance for the general population in the UK is encapsulated in the Eatwell Guide (1). However, this guidance is not considered by some to be sufficiently specific to be applied to older people due to changes in nutritional requirements as a result of ageing (2). The focus on older people is now more relevant due to population ageing across the world (3) as well as in the UK. For example, the number of older people living in England and Wales has increased from 9.2 million (16.4% of the population) in 2011 to 11 million (18.6% of the population) in 2021 indicating the need for a greater focus on the health of older people (4).

Although energy requirements in older people are lower compared to the general adult population due to changes in body composition, requirements for vitamins and minerals remain the same. Additionally, recommendations for protein intake may increase for older adults in order to prevent sarcopenia (5). The consumption of a healthy and nutrient dense diet is important in older people to ensure healthy ageing (2, 3, 6). Physiological and psychological factors associated with ageing such as loss of taste and smell may result in age-related anorexia (7) and reduction in appetite, impacting on older people’ ability to meet their nutritional requirements. This anorexia of ageing can then result in the development of malnutrition (8).

Malnutrition is normally defined as a Body Mass Index below 18.5 kg/m2, or a combination of a BMI of less than 20 kg/m2 along with weight loss (9). It is acknowledged that for older adults a BMI level where malnutrition may exist may be higher than the cut-off for other population groups (9) and malnutrition itself may be associated with obesity (5). Before the Covid-19 pandemic the number of older people living in the community in the UK with malnutrition or undernutrition was estimated to range from between 5 and 10% (10, 11), affecting over 1.3 million people in the UK alone. As well as impacting quality of life in this demographic, this has a significant socioeconomic burden.

Malnutrition was estimated to cost the UK £23.8 billion in 2017 (12)(13). Current societal challenges including the cost of living crisis mean that the number of people living with malnutrition is likely to have increased (14). Data from the Health Survey for England show that 76% of adults aged 65 to 74 years and 73% aged 75 years and over are overweight or living with obesity (15, 16).

Concerted effort is needed to address malnutrition in older people in order to address this public health crisis. Addressing malnutrition requires actions to prevent, recognise and manage malnutrition once identified (7). Part of the effort to address malnutrition with older people include nutritional awareness and literacy(5). There is a need for nutritional education and/or interventions focused on increasing the nutrient density of foods chosen by groups of the older population, including those who are malnourished and/or frail (5). In order to support nutrition strategies to ensure healthy ageing we need to understand where older people source nutrition information, how they understand and respond to information, and how this information can be used to support people to make the dietary changes necessary to meet changing nutritional needs.

### Nutrition education resources used by older people

People are exposed to a range of sources of nutritional information, including media sources (TV, radio, newspapers etc), the internet, as well as labels on food packaging (17). However, nutrition information practices amongst older people (5) have been found to make use of embodied knowledge (that is hidden and unconscious (18, 19)) and we know very little about where older groups of the population source nutritional information, or how this informs what they choose to eat. While there is evidence from those working with this population that current front of pack food labels is a source of nutrition information for older people (20), the population-based approach of providing advice from statutory sources, often focusing on addressing obesity, can have worrying unintended consequences for those who are nutritionally vulnerable.

Nutritional labelling in the UK should comply with regulations set by Government (21), however the current traffic light system is voluntary (22). The aim of nutrition food labelling is to ‘ensure that consumers understand what they are buying and that “it is what it says it is”’ (23). Food labels have been found to be one of the main sources of nutrition information used by the public (24). However, the use of food labels by older people is low (24, 25). Research suggests this group may have difficulties in interpreting the information (25) making them potentially ineffective in facilitating food choices and potentially leading to a worsening nutritional intake. Indeed, there is anecdotal evidence that there may be unintended consequences of the “one-size fits-all” (19) approach to nutritional labelling by those working with older people. The quote below identifies the concern about the inappropriate focus on foods labelled red (high) amongst older people resulting in weight loss and subsequently malnutrition:

> *‘Many of the people attending our services worry about eating things that may be ‘bad’ for them, such as those marked with a danger-invoking, red traffic lights. Sadly, many of these people join our service having lost weight unintentionally, and at risk of, or already malnourished’.* (Sarah Wren, Chief Executive of a social enterprise that supports older people with meals on wheels, nutritional advice, and wider health and wellbeing services).

Maintaining a nutritious, balanced and enjoyable diet plays a crucial role in ageing healthily and avoiding conditions that arise from malnutrition (2). As the world’s population continues to age, understanding what barriers older people face in accessing healthy meals requires more focus. A systematic review undertaken by Host, McMahon, Walton et al. (26) identified that there are a range of factors influencing nutritional intake in older people. The impact of being on the receiving end of nutritional campaigns over a longer period of time as found by Brownie (27) may make decisions about what to eat more challenging.

Although food labels have been found to be one of the main sources of nutrition information their use by older people is low, with research suggesting they may have difficulties in interpreting the information (24, 28). Recognising what messages concerning a healthy diet are being received by older people and where older people find such information in the first place might give us an insight into how decisions about food are being made by this population group.

Although there is research exploring sources of health information in healthy older people (29) and in those with long term conditions (30) as well as sources of nutrition information in the general public (for example (31)), there appears to be limited research exploring sources of nutrition information and their impact on food decisions in older people.

### Aims and objectives

There is no one solution to reducing malnutrition among older people, however, influencing people’s eating habits through provision of nutritional information is worthy of exploration. Obtaining accurate and suitable nutrition information is fundamental to informing healthier dietary choices, positive nutrition attitudes and optimising nutritional status. This scoping review aims to assess the current literature exploring the sources and understanding of nutritional information older people draw on to inform their eating habits and wider notions of a healthy diet.

## Materials and Methods

A scoping literature review following the methodology of Arksey and O’Malley (32) using the checklist developed by Tricco et al (33) was undertaken in order to map the breadth of academic evidence currently available and identify gaps in the research evidence pertaining to nutritional information used by older people. Scoping reviews differ from systematic reviews which focus on answering very specific research questions, rather focusing on assessing and understanding the evidence gaps (34). Peters et al (35) argue that regardless of approach all types of evidence synthesis should be undertaken in a systematic manner and follow methodological guidelines. Therefore, this scoping review was carried out in five stages: identifying the research question, identifying relevant studies, study selection, charting data, and collating, summarising and synthesising results (32). Reporting of this review follows the Preferred Reporting Items for Systematic reviews and Meta-Analysis (PRISMA) extension for Scoping Reviews (34, 35). As is common for scoping reviews we did not conduct a quality assessment of identified studies (36).

### Criteria for inclusion

Inclusion criteria are summarised in table 1. To be eligible for inclusion in this scoping review studies had to include older people aged over 60 years. The review focused on free living older people and does not include studies which focus on particular diseases or long-term medical conditions as the findings would not be generalisable to the wider population (37).

**Table 1.**
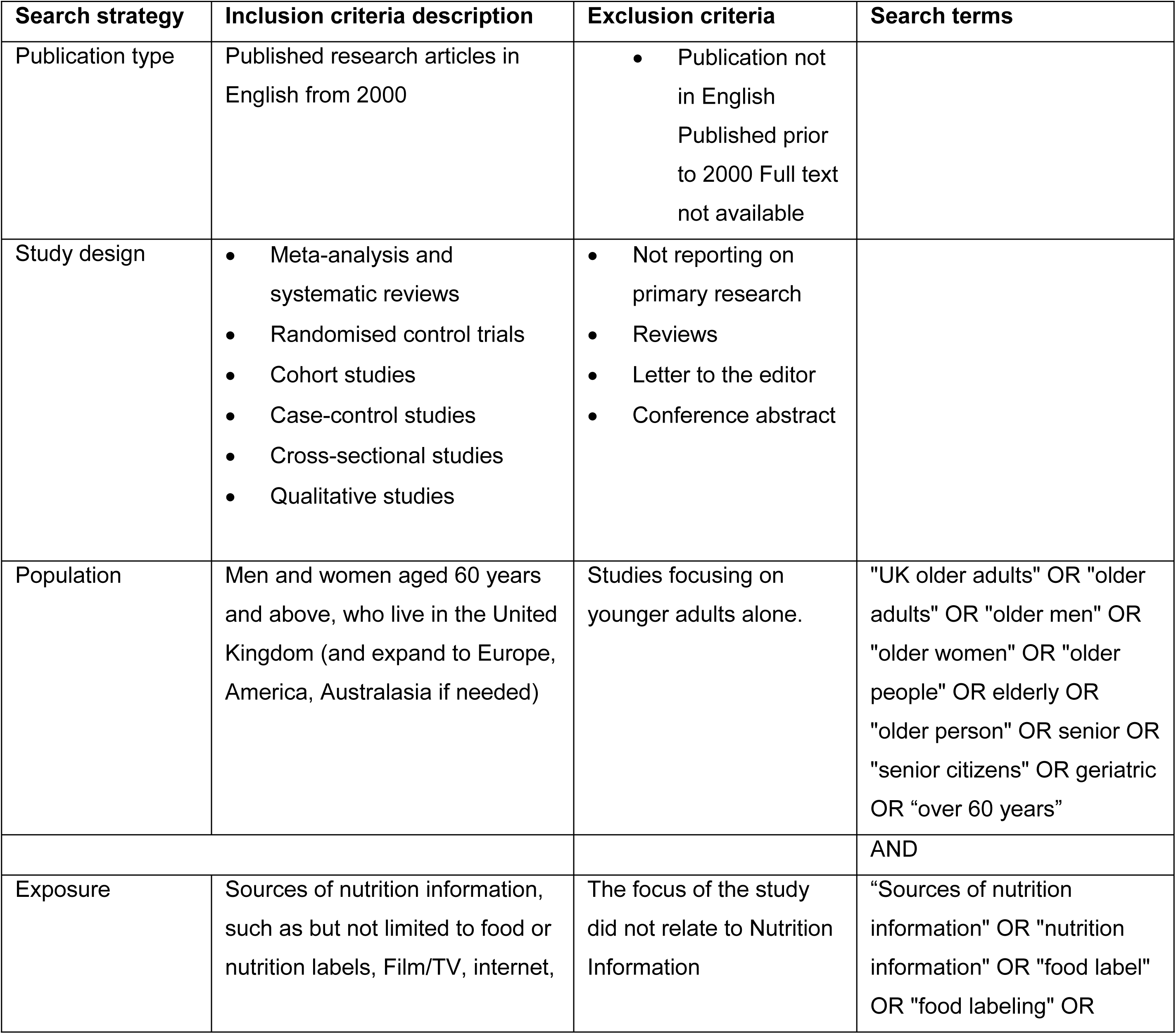
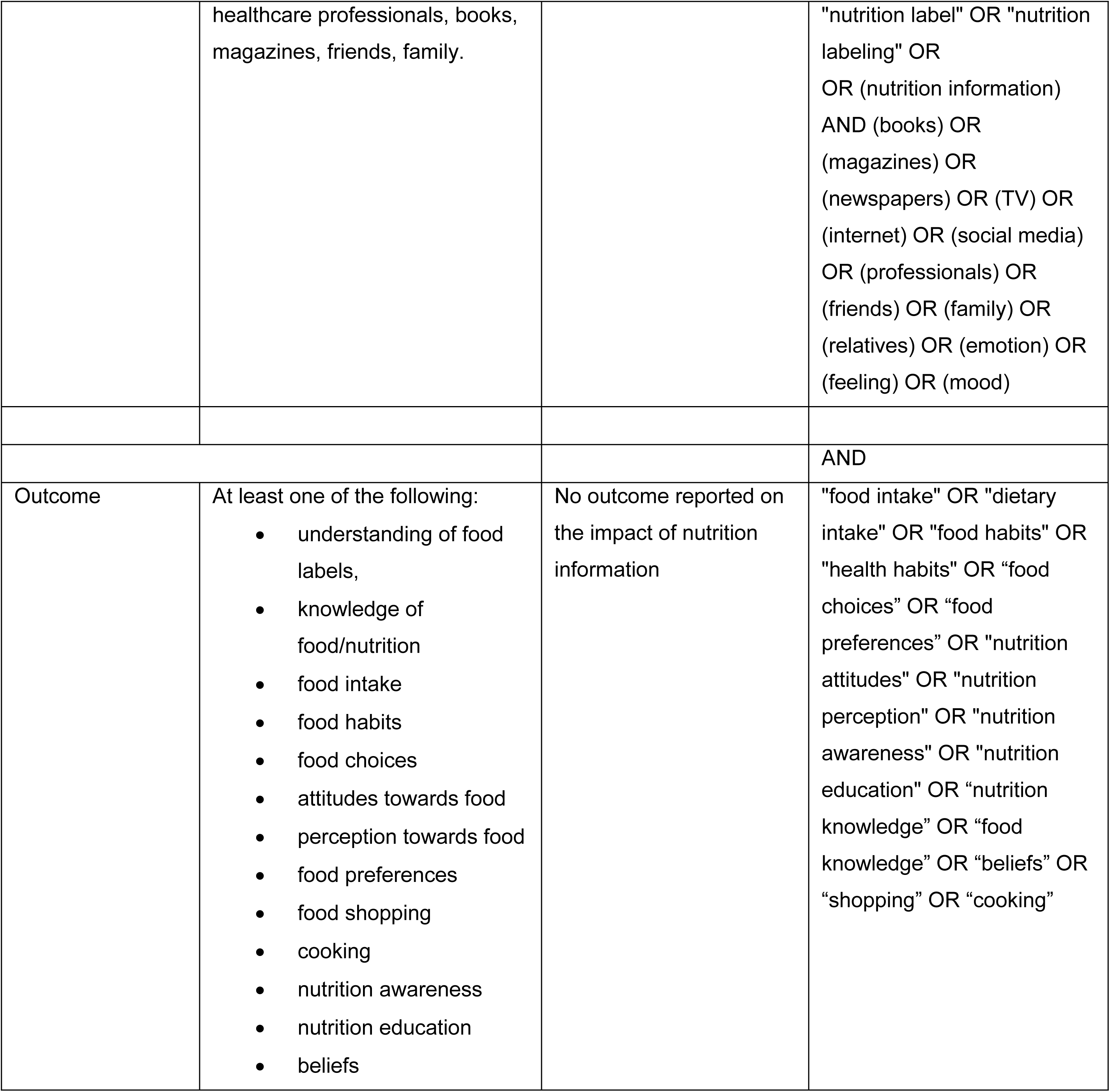
Inclusion criteria description and search terms.

Participants had to have been exposed to or impacted by at least one source of nutrition information. Eligible papers will report outcomes referring to a change in food habits or preferences or a change in the understanding, perception or awareness of nutrition information. Papers were excluded if they focused on younger people alone, did not relate to sources of nutrition information among older people; were reviews, letter to the editor, conference abstract; or if no full text was available. In order that the review reflected current issues, papers were excluded if they were published before 2000. Studies written in languages other than English were excluded.

### Search strategy

The search was conducted in March 2023, across three databases: PUBMED, Scopus and CINAHL. Predefined keywords in the search strategy were based on the Population, Exposure, Outcomes (PEO) framework as outlined in **Table.1**. The search strategies were developed with the assistance of an information manager specialist in the Library and Computing Service at the University of Hertfordshire. Results of the database searches were downloaded into Rayyan screening software, duplicates were removed, and two assessors (TI, EB) independently evaluated the relevancy of each paper against the inclusion/exclusion criteria. A third reviewer (JJ) was asked to settle any differences of opinion. Agreement was reached for all studies included in the final review.

### Data charting process

Data from each study were extracted and charted into excel using the following headings: author, year, aims of the study, country and study setting (for example community or hospital), study design and methods, number, age and gender of participants and finally the outcome of the research and what the study adds to what is known about nutrition information sources used by older people, was extracted, synthesised, and recorded in excel. The studies were then grouped into overall methodology of quantitative, qualitative and mixed methods.

## Results

The initial search on Rayyan yielded 8,864 results, 45 duplicates were identified automatically by Rayyan, leaving 8819 unique entries. Titles and abstracts were screened against the inclusion criteria. No further duplicates were identified. From these results, 31 papers were identified for a full text review. An additional 18 papers were excluded as they failed to meet the inclusion criteria, leaving 13 papers to be included in the final review. Although two papers report on the same study (38, 39), they have both been included as they report on different aspects. The process is summarised in Figure 1 and the studies are summarised in Table 2.

**Figure 1.**
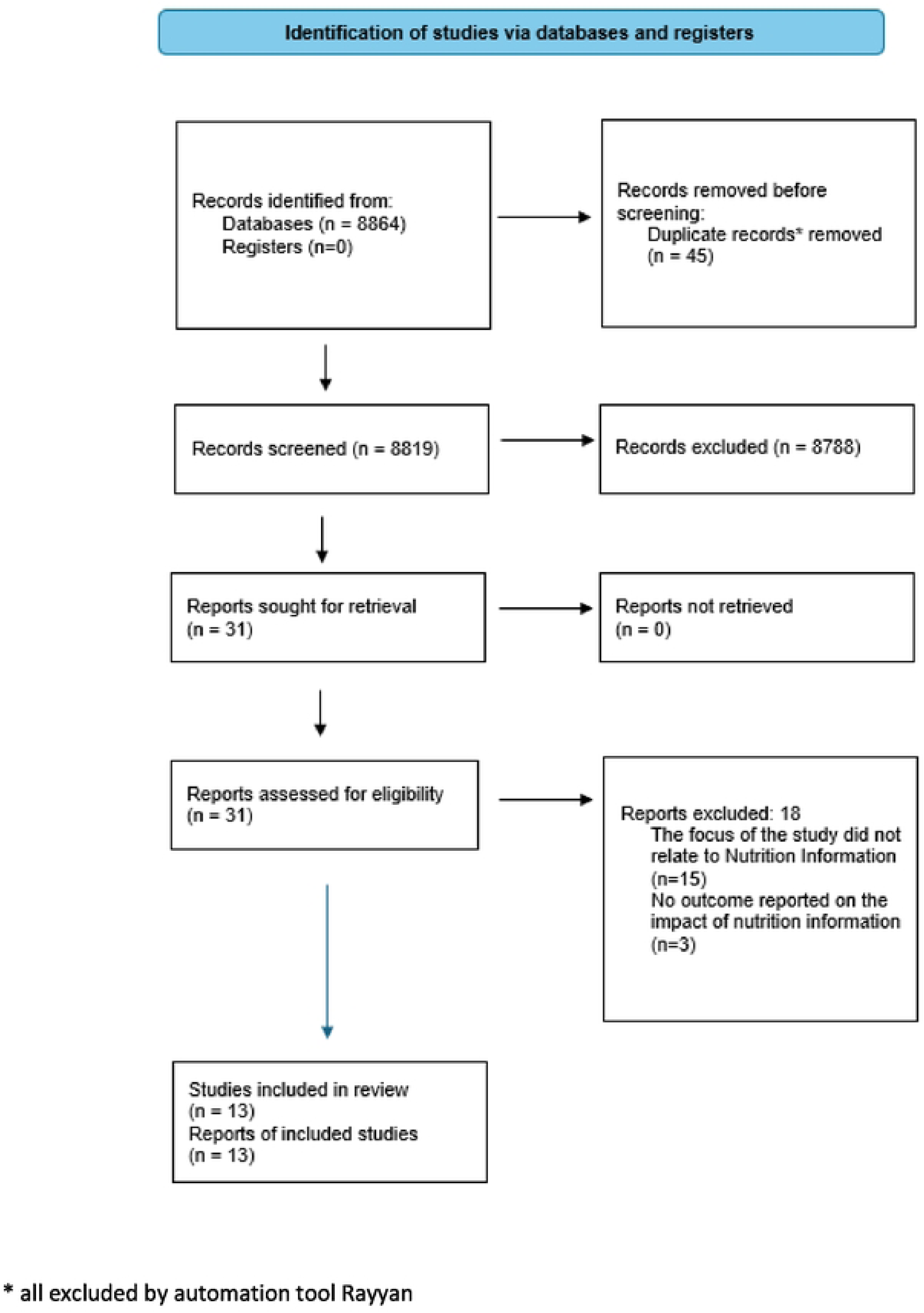
Prisma 2020 flow diagram for the scoping review (adapted from Page, McKenzie, Bossuyt et al. (40)).

**Table 2.**
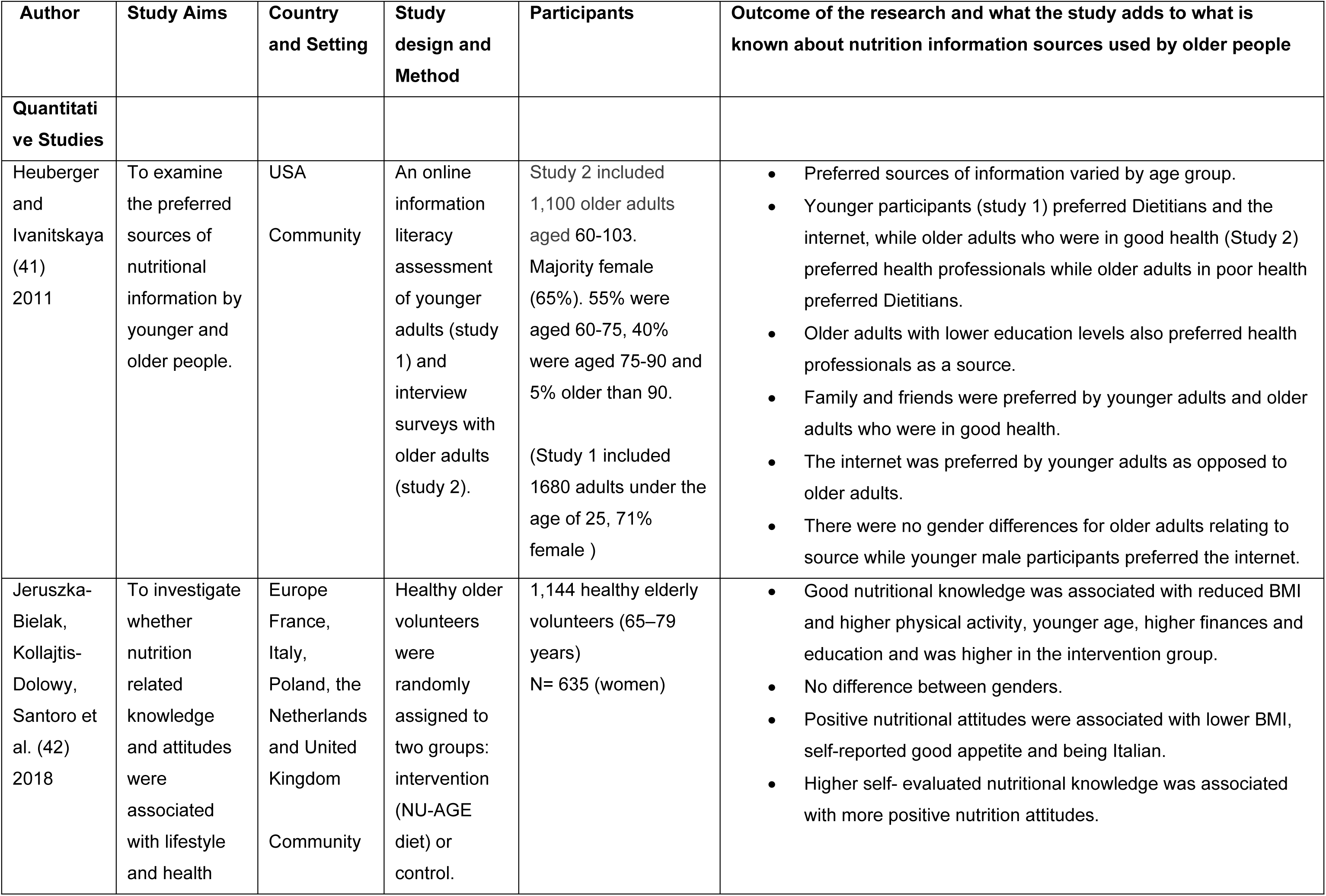
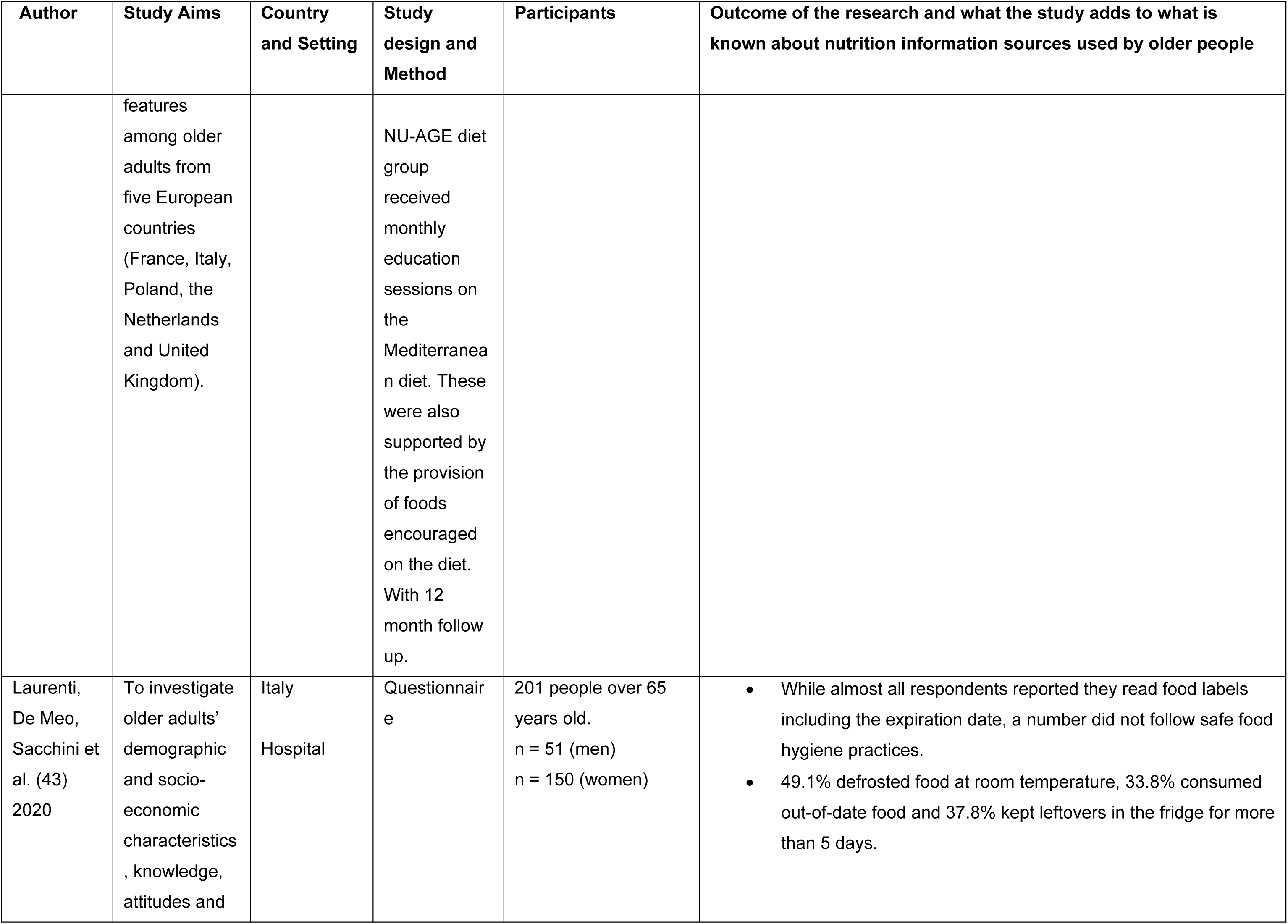
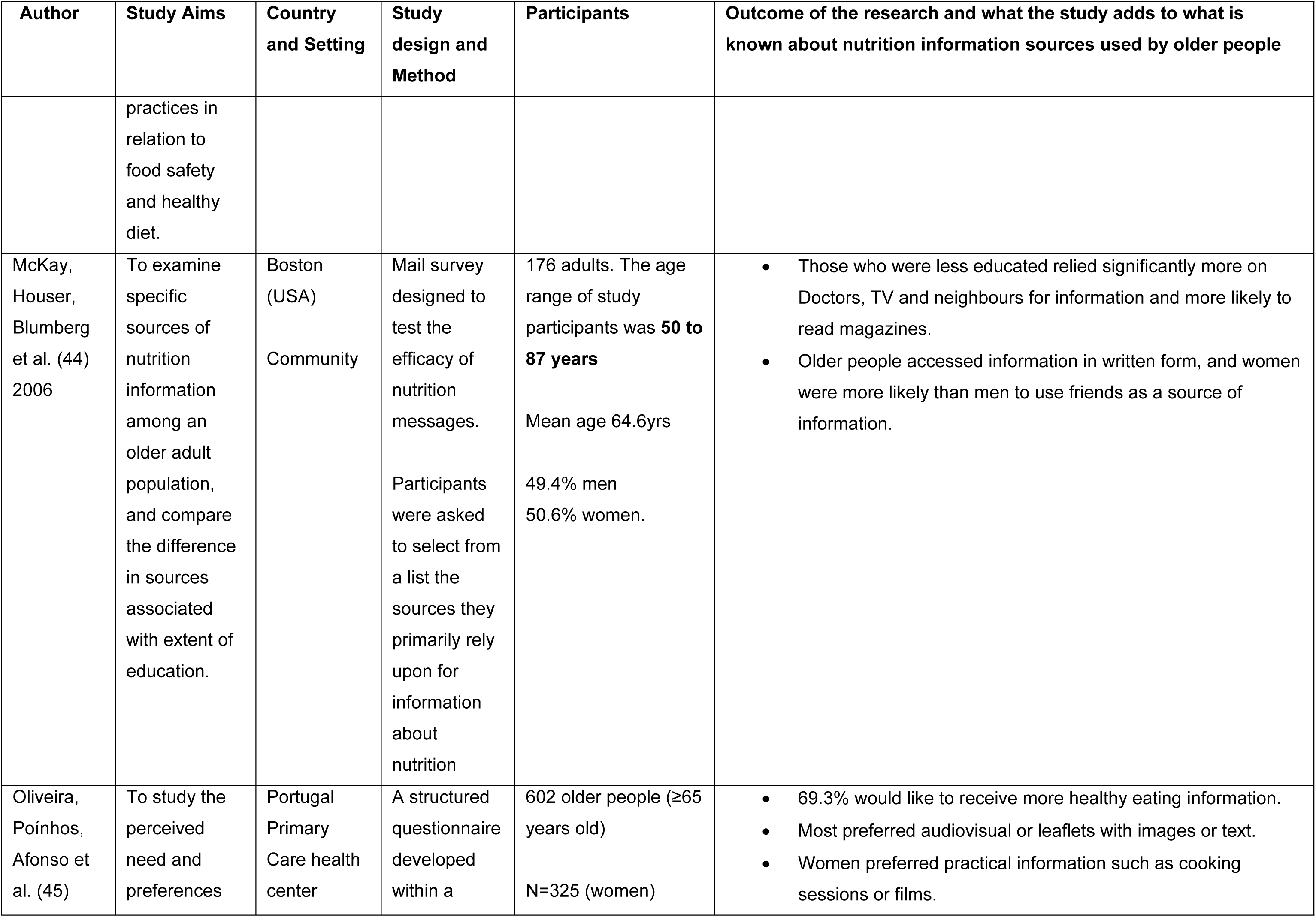
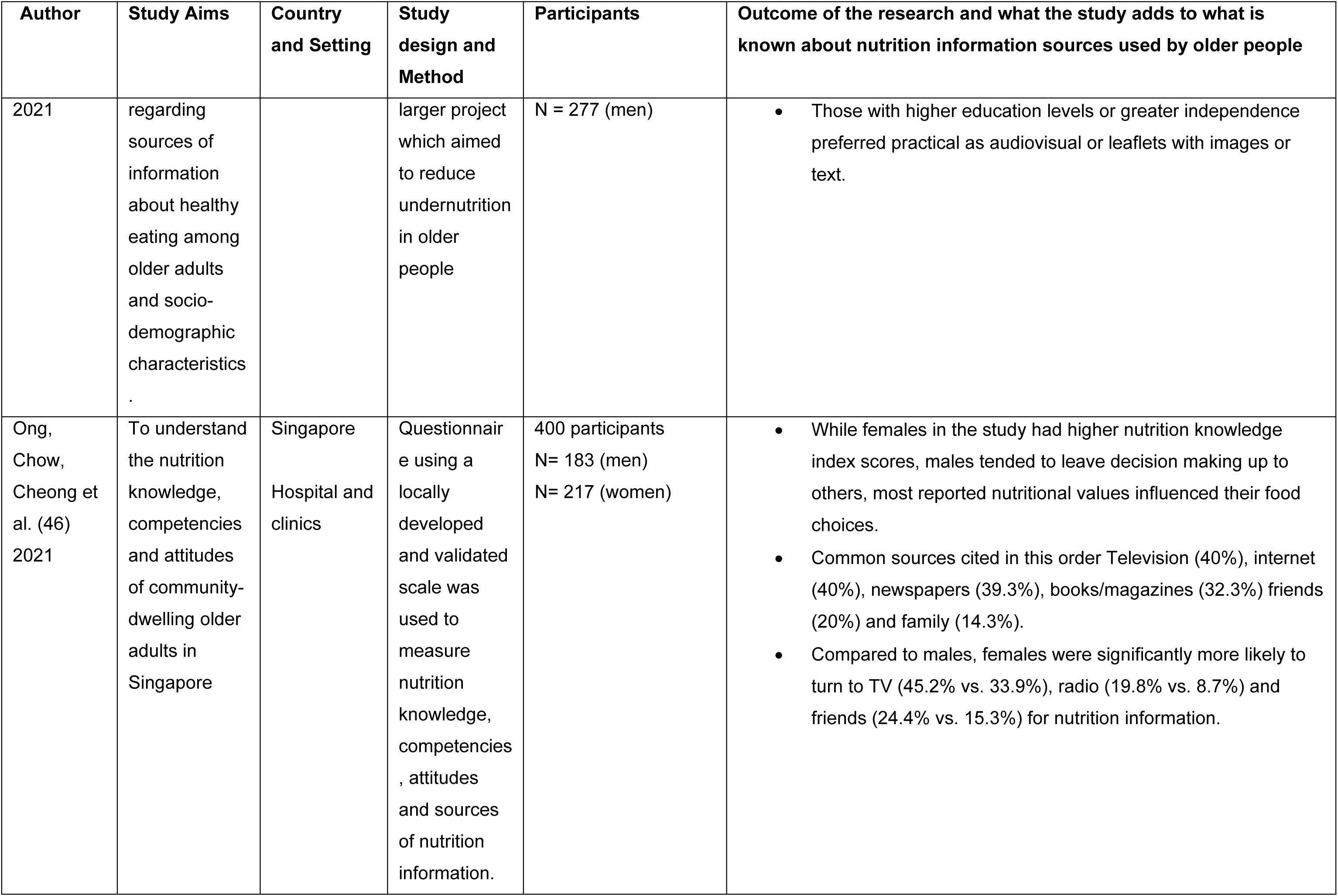
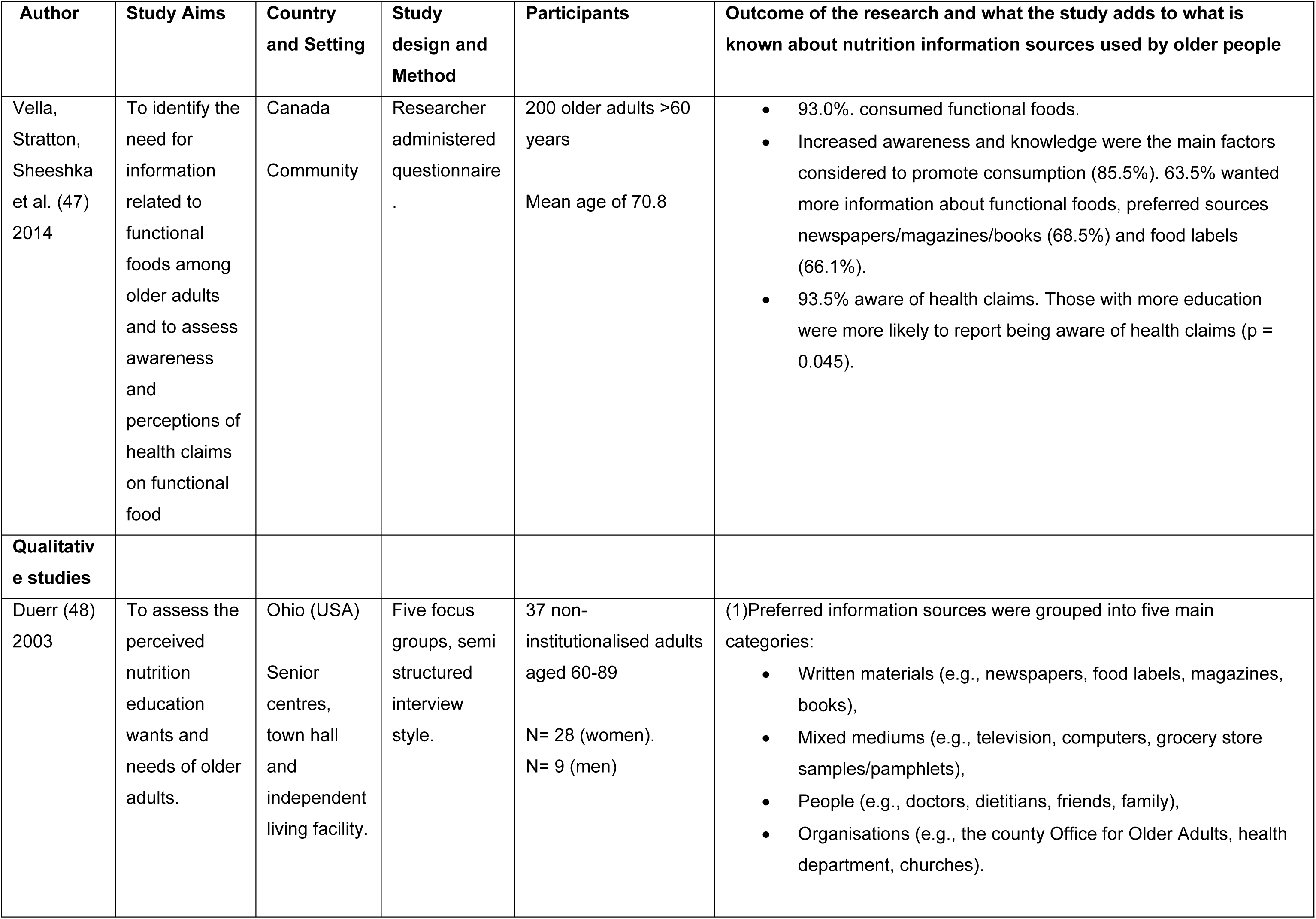
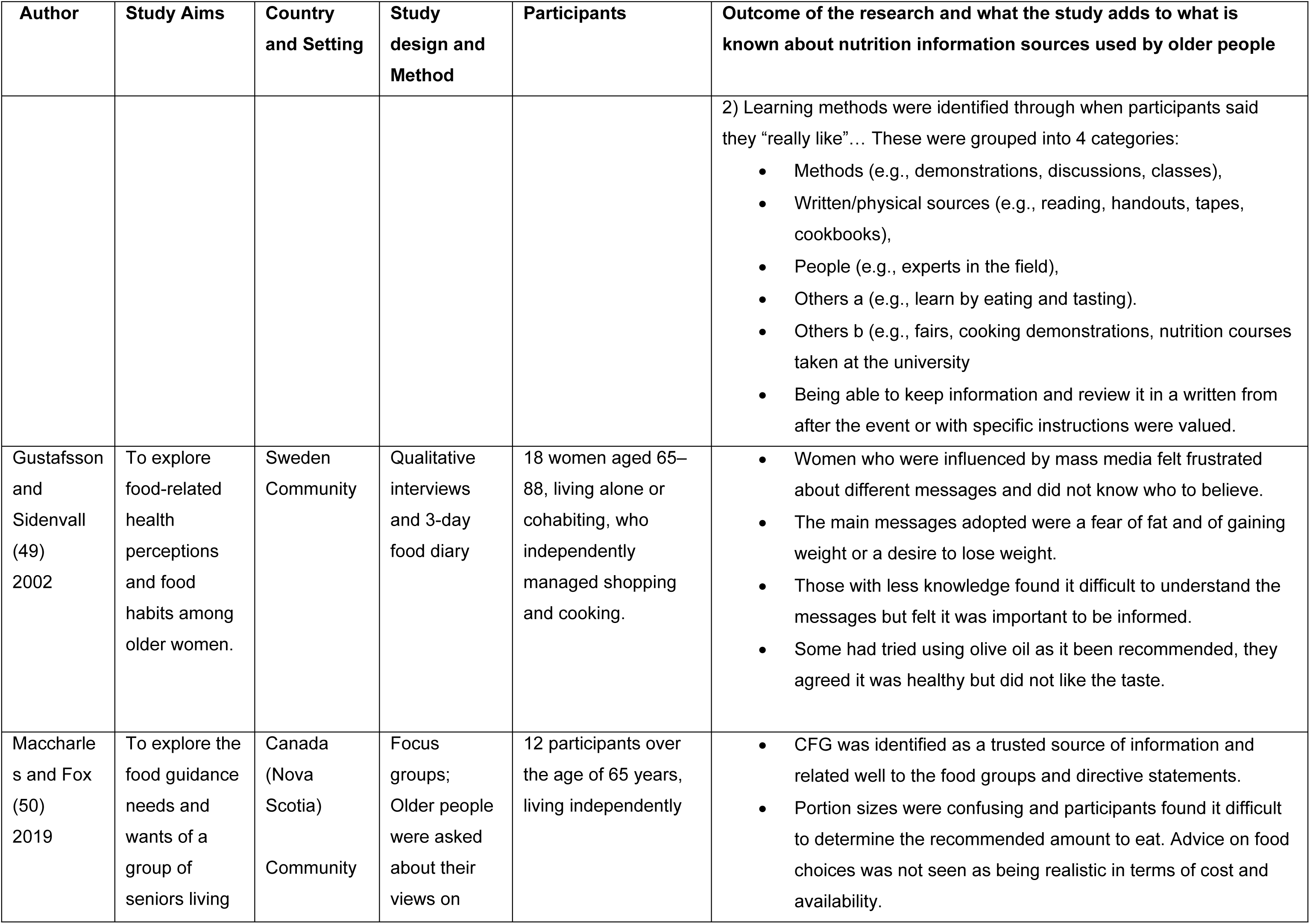
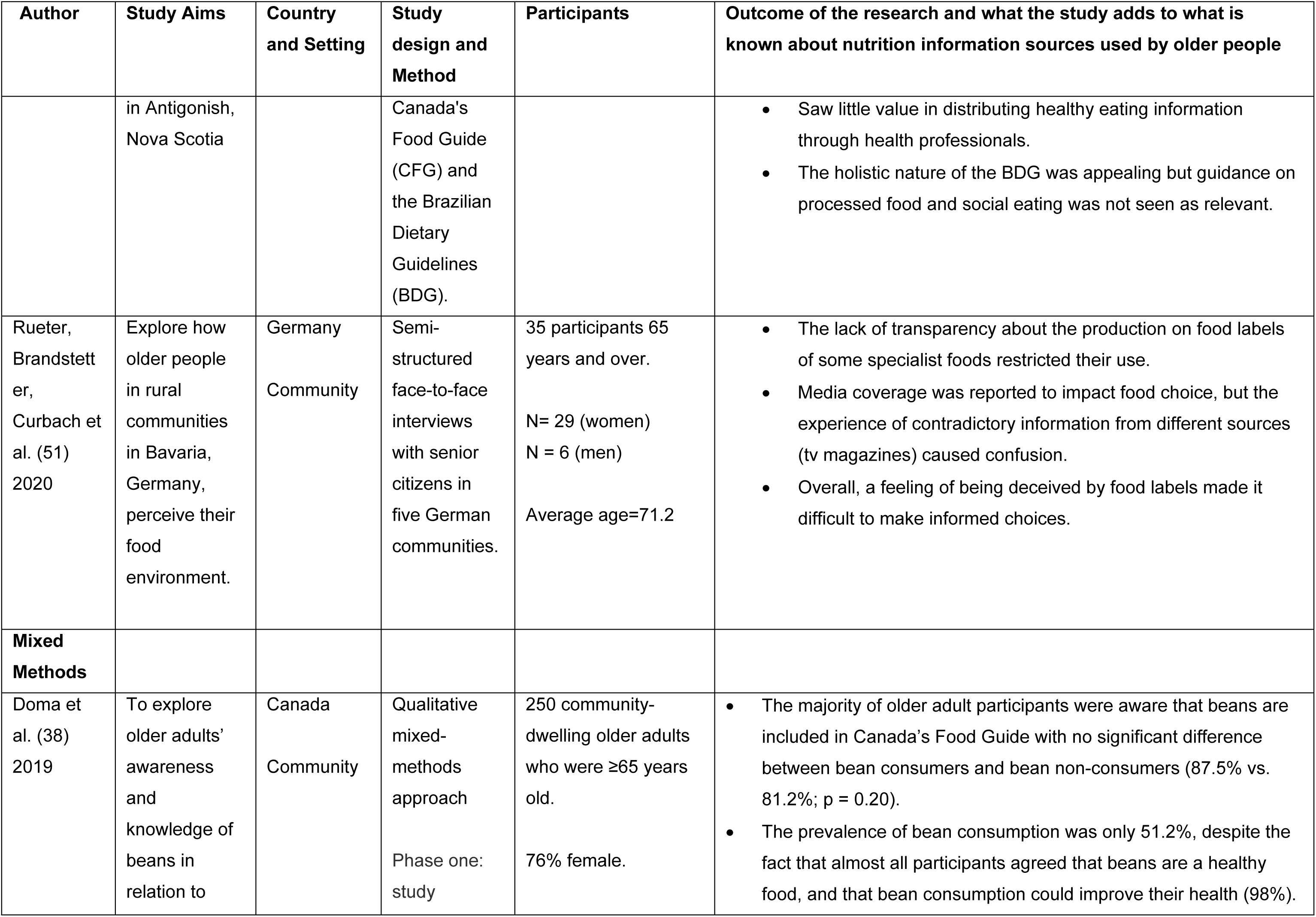
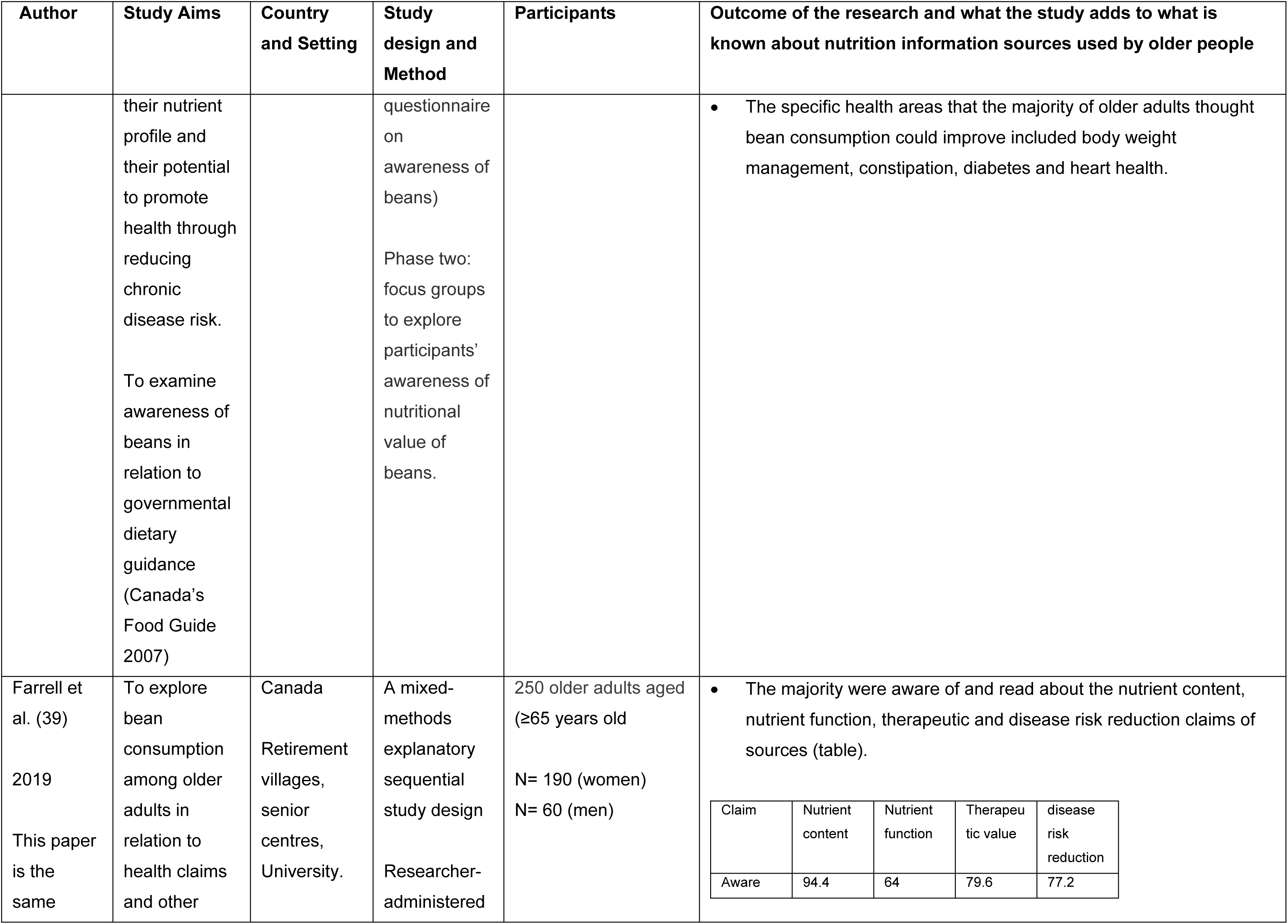
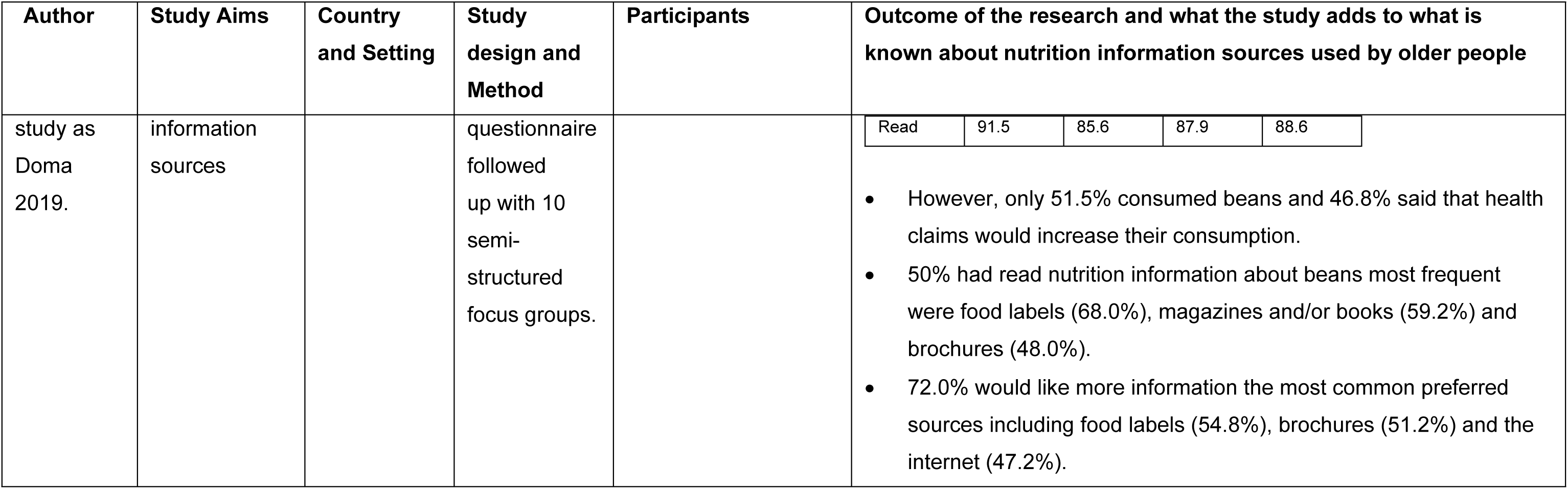
Summary of included studies.

### Characteristics of included studies

#### Study designs

Included papers are comprised of qualitative (*n*=4), quantitative (*n*=7) and mixed method (*n*=2) designs. All of the quantitative papers (41–47) are cross-sectional studies and most record the levels of knowledge or understanding that population groups have regarding nutrition information using survey methods. Only one study involves delivery of an intervention (42). Three of the qualitative papers (48–50) used focus groups and interviews and one study (51) just used interviews to explore outcomes associated with attitudes towards and perceptions of sources of nutrition information. The two mixed method papers (38, 39) combine quantitative methods with further elaboration from participants through focus groups.

#### Settings

Studies were conducted in Canada (*n*=4) (38, 39, 47, 50) 2 of these papers report on the same study (19 and 20), North America (*n*=3) (41, 44, 48), Singapore (46) and European countries of Sweden (*n*=1) (49), Portugal (*n*=1) (45), Italy (*n*=1) (43) and Germany (*n*=1) (51), one study (42) was undertaken across five different European countries (France, Italy, Poland, the Netherlands and United Kingdom (22% of the 1144 participants were from the UK).

The majority of studies recruited participants from community settings (*n*=6) (41, 42, 44, 47, 49, 50) including centres for older people such as town halls or retirement villages (*n*=4) (38, 39, 48, 51), while three recruited participants from hospitals (*n*=2) (43, 46) and healthcare centres (45).

#### Participants

##### Age

The age range of participants across the studies was 50-103 years. Although the inclusion criteria for studies to involve adults aged over 60, one study that involved people aged between 50-87 was included as the mean age of participants was 64.6 years.

##### Gender

In one study (49) all participants were women. There were no studies where all participants were men. One study (44) included 50.6% women and 49.4% men. The overall distribution of participants between women and men was 61% women 39% men. This may be because prior research has found that women have often been found to have more interest in food-related issues so are more likely to take part in studies. Also that they may have more nutrition knowledge than men and so may feel more confident in taking part in research about this topic (52).

### Outcomes of studies

In this section we explore the main themes identified through the analysis of the studies. The main themes as follows are listed by study in table 3 below: Need for information; Embodied nutritional knowledge and impact on food choice; Impact of sources of nutrition information on food practices; Preferred sources; Gender differences and sources of nutrition information; Education levels and sources of nutrition information; Trust and conflicting messages.

**Table 3.**
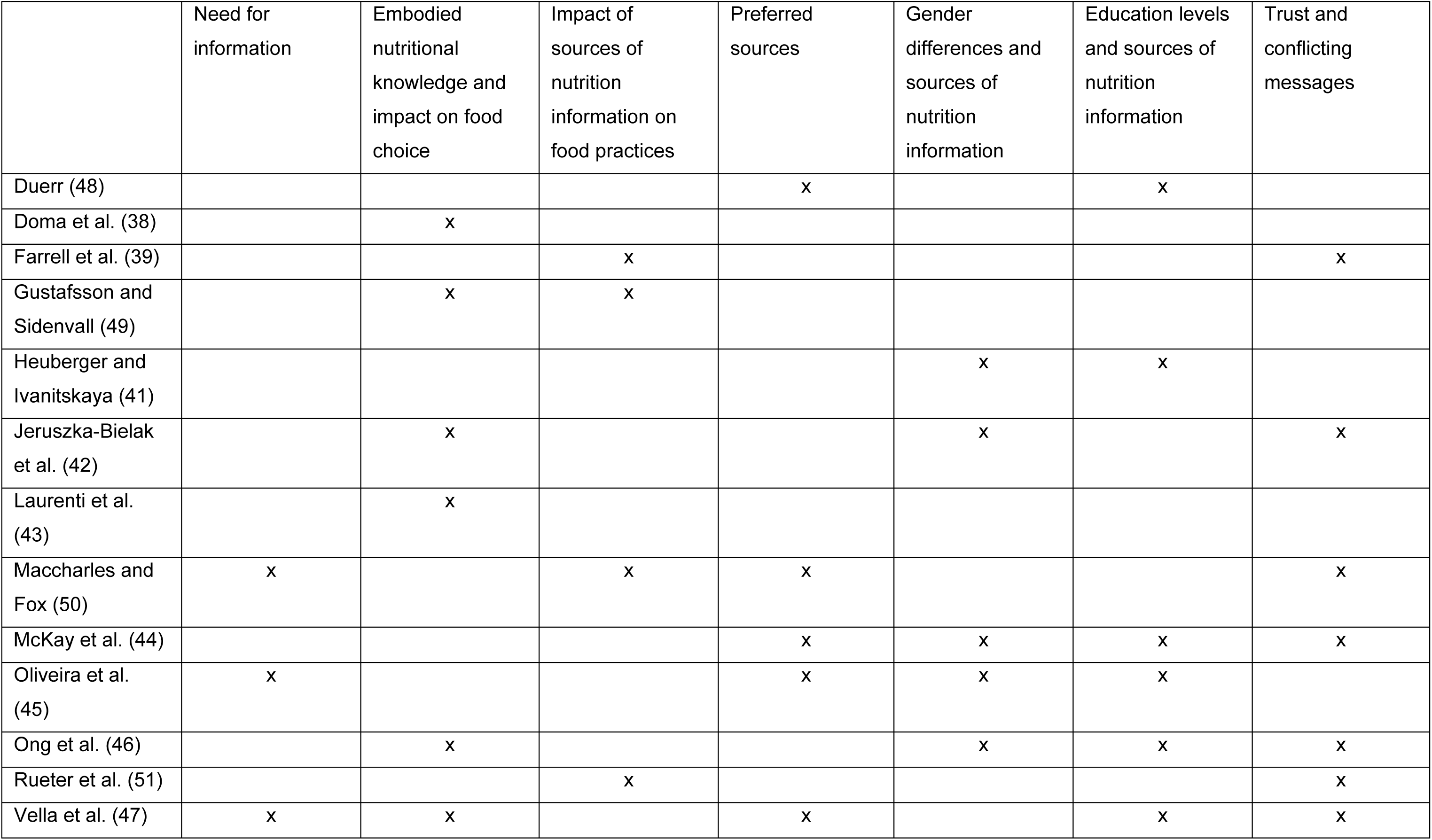
Studies by theme.

### Sources of nutritional information

The studies found that a wide range of sources of nutritional information were used by older people. The most common sources were magazines which were identified by eight studies (39, 42–44, 46–48, 51) ; television (six studies) (41, 43, 44, 46, 48, 51); dietitians (six studies) (39, 41–43, 46, 48) and family and friends (six studies) (39, 41, 44, 46, 48, 51). These are summarised in table 4.

**Table 4.** Collation of sources of information identified by the studies.

|  | Magazines | Dietitian | Embodied Knowledge | Family and Friends | GPs/doctors | Television | Books, recipe books | Food labels | Internet | HCPs | Newspapers | Leaflets booklets | Radio | Audio/audiovisual | Health authorities | National Food guide | Practical, courses | Church | Mass media | Nurse | Nutritionists | Organisations | Posters | Scientists |
| --- | --- | --- | --- | --- | --- | --- | --- | --- | --- | --- | --- | --- | --- | --- | --- | --- | --- | --- | --- | --- | --- | --- | --- | --- |
| Duerr (48) | x | x |  | x | x | x | x | x | x |  | x | x |  | x | x |  | x | x |  |  |  | x |  |  |
| Doma et al. (38) |  |  | x |  |  |  |  |  |  |  |  |  |  |  |  | x |  |  |  |  |  |  |  |  |
| Farrell et al. (39) | x | x |  | x | x |  | x | x | x |  |  |  |  |  |  |  |  |  |  |  |  |  |  |  |
| Gustafsson and Sidenvall (49) |  |  |  | x |  |  |  |  |  |  |  |  |  |  | x |  |  |  | x |  |  |  |  | x |
| Heuberger and Ivanitskaya (41) |  | x |  | x |  | x |  |  |  | x |  | x | x |  |  |  |  |  |  |  |  |  |  |  |
| Jeruszka-Bielak et al. (42) | x | x | x |  | x |  | x | x | x | x |  |  |  |  |  |  |  |  |  |  |  |  |  |  |
| Laurenti et al. (43) | x | x | x |  | x | x |  |  | x | x |  |  |  |  |  |  |  |  |  |  | x |  |  |  |
| Maccharles and Fox (50) |  |  |  |  |  |  |  |  |  |  |  |  |  |  |  | x |  |  |  |  |  |  |  |  |
| McKay et al. (44) | x |  |  | x | x | x |  |  |  |  | x |  | x |  |  |  |  |  |  |  |  |  |  |  |
| Oliveira et al. (45) |  |  |  |  |  |  |  |  |  |  |  | x |  | x |  |  | x |  |  |  |  |  | x |  |
| Ong et al. (46) | x | x | x | x | x | x | x |  | x |  | x |  | x |  |  |  |  |  |  | x |  |  |  |  |
| Rueter et al. (51) | x |  |  | x |  | x |  | x |  | x |  |  |  |  |  |  |  |  |  |  |  |  |  |  |
| Vella et al. (47) | x |  | x |  |  |  | x | x |  |  | x |  |  |  |  |  |  |  |  |  |  |  |  |  |
| Number of studies | 8 | 6 | 6 | 6 | 6 | 6 | 5 | 5 | 5 | 4 | 4 | 3 | 3 | 2 | 2 | 2 | 2 | 1 | 1 | 1 | 1 | 1 | 1 | 1 |

### Need for information

Three studies exposed the need for nutrition information. Oliveira et al. (45) who undertook a questionnaire with 602 older people in Portugal found that most participants were concerned about healthy eating (87.5%) and would like more information (69.3%). Similarly, Vella et al. (47) found that their participants wanted more information about functional foods. However, the need for nutrition information may be impacted by the perceived relevancy of the information. For example Maccharles and Fox (50) who explored the Canada Food Guide with a small group of older people in focus groups (women n=10, men n=2) found that information that was related to daily living and eating was not considered relevant or a needed part of the Food Guide.

### Embodied nutritional knowledge and impact on food choice

The studies appear to indicate that the relationship between embodied nutritional knowledge (19) and food chosen/eaten is not straightforward. Some studies found an association between nutritional knowledge and eating practices, for example identifying embodied information i.e. knowledge held by participants as a source of information that impacted on food choice. For example, Vella et al. (47) found that knowledge already possessed by participants promoted the consumption of functional foods. and Gustafsson and Sidenvall (49) and Ong et al. (46) (amongst men) found that lack of knowledge impacted on being able to interpret health information and decision making on food choices. However, Doma et al. (38) who undertook a questionnaire with 250 older people followed by focus groups with 10 participants found that the possession of knowledge about the healthfulness of beans did not result in increased consumption by participants.

The potential positive impact of nutritional knowledge was explored in an intervention study which delivered nutritional information sessions, including how to use food labels to make healthy food choices, to older people with a 12 month follow up Jeruszka-Bielak et al. (42)). They found increased nutritional knowledge had a statistically significant positive impact on health outcomes such as a lower BMI, higher physical activity and positive nutrition-related attitudes. They explored this further with the impact of new knowledge gained through monthly education sessions on the Mediterranean diet with 1144 healthy older people from 5 European countries including the UK. (The intervention was supported by the provision of foods encouraged on the diet). They found that the intervention improved knowledge and positive attitudes towards healthy eating compared to the control group. However, this link between perceived knowledge and BMI is at variance with the study undertaken by Laurenti et al. (43) who undertook a self-administered questionnaire with 201 older people (women n=150, men n= 51) who were attending a ward-based, outpatient clinic gym session for older people in Italy. They found that while 64.2% of participants believed that they followed a balanced diet, 51.2% were either overweight or obese.

### Impact of sources of nutrition information on food practices

Studies exposed the different impact that nutrition information had on food choice. Rueter et al. (51) who sought to explore factors that influenced food choice with 35 older people through semi-structured interviews found that although endorsement of specific foods in mass-media, specifically television and magazines, has the potential to influence food choice, participants experienced contradictory information and a feeling of being deceived by information on food labels. Gustafsson and Sidenvall (49) whose aim was to explore food related health perceptions with a small group of 18 older women also found frustration with different messages, and lacking the skills to be able to critically appraise advice, they were not sure who to believe. Similarly, Maccharles and Fox (50)￼ also with a small group of participants (n=12) found that the portion size information in the Canada Food Guidelines was confusing. However Farrell et al. (39)￼￼who explored the link between health claims and bean consumption in 250 older people also in Canada found that health claims on food labels were the most commonly accessed and preferred source of information about the health benefits of beans.

### Preferred nutrition information sources

Some studies examined where older people preferred to source nutritional advice. For example, McKay et al. (44) undertook a survey with 176 older people (mean age 64.6, 50.6% women, 49.4% men) and identified that the sources they used for information were doctors (61.8%), newspapers (61.8), magazines (60.1%) and television (49.1%) with level of education affecting the type of print media they accessed. Vella et al. (47) explored sources of information about functional foods with 200 older people (70% women) through a researcher completed questionnaire and found that 68.5% preferred newspapers, magazines and books as sources of information about functional foods. Maccharles and Fox (50) found while Government sources such as food guidelines were trusted, participants felt that HCPs were not a useful source of healthy eating information. Duerr (48) used focus groups to explore information sources currently used and what learning methods were ‘liked’ by participants with 37 older people. This study was able to expose how participants processed information (for example by taking information home, reading it through and saving for later use). They identified the value of practical learning methods such as demonstrations, discussions, classes, eating and tasting and the importance of being given practical information on how to adapt recipes to make them healthier. Similarly Oliveira et al. (45) who undertook a questionnaire as part of a larger study aiming to reduce undernutrition in older people found that practical sources of information were a preference.

### Gender differences and sources of nutrition information

Although Heuberger and Ivanitskaya (41) who undertook a survey with 1100 older people (65% women) in the USA did not find differences in preferred sources of nutrition information between men and women, a number of other studies did. Oliveira et al. (45) (54% women, 46% men) found that a higher proportion of women preferred to receive information through practical cooking sessions (38.5%) than men (28.6%) (p =0.044), while Ong et al. (46) found that women were more likely to use television, radio and friends as a source of nutrition information, similarly McKay et al. (44) who undertook a survey in Boston with 176 older people (50.6% women and 49.4% men) also found that women relied on friends more often than men. However, this was in contrast to Jeruszka-Bielak et al. (42) who found that 39% of men preferred to make use of friends and family versus 33% of women.. This study also found that more women than men preferred books and magazines (61% vs 49%), while more men preferred internet (43% vs 35%).

### Education levels and sources of nutrition information

Educational level impacted on preferred sources and use of information. For example Oliveira et al. (45) found that those with a higher educational level preferred leaflets with text and audiovisual materials (p values 0.013, 0.027 respectively). While McKay et al. (44) found that those with more than 4 years of college education (n=92) were more likely to use the New York Times, Time, Newsweek and national public radio. However, those with less than 4 years of college education (n=84) were less likely to use magazines (specifically Good Housekeeping), neighbours and doctors, (although the figures are small). Conversely Heuberger and Ivanitskaya (41) found that a higher percentage of those with a lower level of education were more likely to refer to a professional source of nutrition information than those with a higher level of education (<12 years 73% of women, 74% of men, versus >16 years 67% of both women and men). Vella et al. (47) found that those with a higher education level were more likely to be aware of food claims on food labels, similarly Ong et al. (46) found that a higher educational level enabled the ability to critically appraise information about nutrition.

### Trust and conflicting messages

Questions of credibility and trust in sources of information were noted. Seven studies identified the high level of trust of information and advice provided by health professionals (39, 42, 47, 48, 50, 51). However, two studies (47, 48) explored the sense of mistrust that older people had in the current food system and health claims that they have been exposed to.

The navigation of conflicting or incompatible nutrition information was also identified. For example McKay et al. (44) found that some older people perceived inconsistencies in the health claims and advice found across different sources and found this discordance confusing when trying to follow a healthy diet. Ong et al. (46) highlighted the conflict between traditional practices or beliefs that had been instilled since childhood and current nutrition guidance. Trust in a nutrition information source would encourage its use by older people, however mistrust may prevent use of the nutrition information source and in the food that the source may be promoting.

## Discussion

We believe this is the first scoping review bringing together academic literature focusing on nutrition information and older people. The review found that older people used a wide range of information sources, including food labels, written media, health care professionals and family and friends and identified the positive ongoing impact of a nutrition education intervention on food practices. Use of nutritional information appears to be mainly affected by educational level but there are also gender differences with women appearing to be more engaged with nutrition information. The source of nutrition information was important and affected levels of trust and needed to be relevant and accessible. The review also found a potential positive impact of practical delivery of nutrition education information through sessions involving recipe adaptation, cooking and tasting the food.

Most of the studies were undertaken outside the UK, relying on questionnaires to collect data. One study included older people in the UK and involved an intervention (42).

The wide range of sources of nutrition information identified in this current study was in common with other studies exploring both health and nutrition information practices (30, 31, 53). Other studies have found high reliance on information provided by health care professionals exploring health information practices in older people and in people with long term conditions. For example Hurst (29) undertook an exploratory qualitative study with older people and found that the impact of age and the development of health-related conditions meant that healthcare professionals were the most common source of health information. The findings relating to credibility and trust in information provided by dietitians and doctors are supported by other academic work that illustrates the high levels of confidence older populations have in them (54). However, research suggests that this trust and therefore following advice from health care professionals may be outweighed by older people’s preferences and beliefs about what foods are good for them (55). Also Gustafsson and Sidenvall (49) note that, while mistrusting sources, older people did not feel as though they possessed the necessary skills or knowledge to critically appraise the information they came across.

Research has found food label use amongst older people to be low (24). In this current review the value of food labels was found to be conflicting. Although the older people in Farrell et al. (39) found them useful for highlighting health claims about the importance of beans, Rueter et al. (51) noted that their participants were concerned about the potential for food labels to deceive.

Magazines and television were common preferred sources identified in the studies. There is limited research identifying magazines as a specific source of nutrition information, however Wills, Dickinson, Short et al. (56) found that as a source magazines may not contain robust evidence-based information. The value of television as a potential source of information was found by Rivero-Jiménez et al. (55), where participants in isolated rural communities in Spain relied upon television as a source of social interaction.

The internet was a preferred source by participants in 5 of the studies featuring either second or third in preferences where this was scored, however the internet had a lower preference level where older people’s preferences were compared with younger people’s preferences (41). Although the internet is often featured as a main source of information in people living with long term conditions such as type 2 diabetes (T2DM) (30) research has found that the internet is more often made use of by younger people living with T2DM (57, 58). Studies exploring sources of nutrition information have also found that the internet featured as a common source. For example, Ruani et al. (31) found that members of an organisation aimed at those with an interest in learning about nutrition, identified nutrition websites and google searches as common sources.

However, they also identified nutrition related books as a preferred source,a result that is in line with the findings of this review.

Rueter et al. (51) (identified in this current study) found that information found in the mass media was conflicting and confusing. Other works have also pointed to older people’s perceptions of health claims as being used as marketing tools, rather than being credible and reliable sources of nutrition information (59). Elsewhere, it has been highlighted that such feelings of distrust have health implications and can affect consumers’ abilities to maintain a healthy and balanced diet (60). Understanding the health effects of mistrust amongst older consumers might provide valuable insights into further understanding the impact of nutrition information or lack of it on the causes of malnutrition.

### Further research and implications

The scoping review has identified a need for further research in this area, particularly in the UK. Future work could examine the role of, and advice given to older people, about food and nutrition by healthcare professionals, as they were often a preferred and trusted source of nutritional information. In order to identify the impact of sources on older people, research should include a greater focus on information received on food choice.

The study findings will be of value particularly to nutrition and dietetic professionals working with older people. Opportunities to assist in interpreting nutrition information sources such as food labels and exploring views and opinions about nutrition information accessed by older people should be taken during consultations or nutrition communication events.

### Conclusion

Overall, this scoping review has identified the wide range of sources used by older people, that the usability of these sources is impacted by levels of education and by trust, and the potential for health claims on food labels to communicate nutrition information. There are opportunities for further research to explore the impact of nutrition information on food choice and for health care professionals particularly nutritionists and dietitians to support older people in being able to make use of nutrition information in order to make healthy food choices to promote heathy ageing.

## Data Availability

All relevant data are within the manuscript and its Supporting Information files.

## Acknowledgements

This work was supported by the Biotechnology and Biological Sciences Research Council Food for added life years: Putting research into action (Food4Years) grant. [Award Reference RCP1007006, Funder Reference BB/W018349/1]. All authors contributed to the study design and agreed the final article. JM undertook the analysis and constructed the final article with AD. TI and EB extracted the data and JJ reviewed the data and confirmed study inclusion. Dr Rosalind Fallaize is acknolwedge for their support in the design of the search process.

